# A global cross-sectional survey of health professionals’ interest-confidence gaps in value-based health care implementation: a learning needs assessment

**DOI:** 10.64898/2026.06.10.26355253

**Authors:** Sally Lewis, Alice O Andrews, Hamish Laing

## Abstract

**Objectives:** Value-Based Health Care (VBHC) increasingly guides health system redesign internationally. Despite the increasing availability of VBHC education, gaps remain between health professionals’ conceptual understanding of VBHC and their confidence to implement it in practice. This study assessed perceived learning needs and preferences of healthcare professionals across foundational topics essential to VBHC implementation.

**Design:** Cross-sectional online survey study

**Setting and participants:** The survey was distributed to the global VBHC community and yielded 518 responses. Most respondents were based in the UK and Ireland (51%) and 65% had more than 10 years of experience in the health sector. Participants represented a variety of professional backgrounds, including clinicians (34%), operational or executive managers and leaders (22%), and life sciences or procurement professionals (13%).

**Primary and secondary outcome measures:** Primary outcome measures included self-reported interest and confidence across 15 VBHC domains and the magnitude of the gap between them. Secondary outcomes included perceived implementation challenges and preferred VBHC learning approaches, including prior engagement with VBHC-related learning.

**Results:** Respondents identified substantial VBHC implementation challenges, including implementing outcome measurement (62.4%), conflicting priorities (57.7%), and resistance to change (56.8%). Interest in all VBHC domains was high (median >= 80/10), while confidence to implement remained substantially lower across most domains (median ti=50/100). The largest interest-confidence gaps were observed for reimbursement mechanisms, costing methodology, and overcoming implementation challenges. Interactive learning approaches, including in-person seminars/workshops (55.2%) and online masterclasses (53.9%) were preferred over self-directed formats.

**Conclusions:** This international survey identified consistent gaps between health professionals’ interest in VBHC and their confidence to implement key VBHC domains in practice. Addressing these gaps through advanced, targeted and contextual education may support more effective and sustainable VBHC implementation in practice.

## INTRODUCTION

Since the publication of Porter and Teisberg’s seminal book “Redefining Healthcare(1),” Value-Based Health Care (VBHC) increasingly has influenced policy, service redesign, and professional discourse across health systems internationally. Implementing the principles of VBHC is a substantial undertaking, requiring technical and cultural change, robust data infrastructure and a will to redesign models of care to improve outcomes and ensure sustainability. As VBHC principles have been introduced across diverse settings, persistent practical barriers have emerged. Their continued presence suggests that despite the availability and global uptake of executive education and master’s programmes, important gaps remain between conceptual understanding and the practical capability required for implementation.

Feedback from participants in executive education programmes has highlighted demand for more advanced and applied learning to support professionals in overcoming these barriers. We sought therefore to better understand the magnitude and nature of skill/capability gaps, as well as the learning needs and preferences of the global healthcare and health policy workforce. To this end, we developed an exploratory Learning Needs Assessment survey examining respondents’ interests in the key VBHC domains and their self-reported confidence to implement these elements in routine practice.

## MATERIALS and METHODS

### Study design and study population

An exploratory, cross-sectional Learning Needs Assessment questionnaire was distributed electronically through a shareable Qualtrics^™^ link to organisations and individuals involved in VBHC worldwide, using a two-stage, non-probability sampling strategy to maximise reach and response rates (see Appendix A). The study complied with the UK and European Union’s General Data Protection Regulations (GDPR) and was approved by the research ethics board at Swansea University. No personally identifiable information was collected, and IP addresses were not retained in the analysis. The first page of the survey included an informed consent statement explaining that participation was voluntary, and that submission of the survey indicated consent for use of the anonymised, aggregated data in research and publication. This study is reported in accordance with the Strengthening the Reporting of Observational Studies in Epidemiology (STROBE) guidelines.

### Instrument

The Learning Needs Assessment questionnaire was developed to assess educational needs, priority topic areas, and preferred formats for advanced educational programmes tailored to professionals interested in VBHC. The questions were derived from a review of the literature (2,3), alumni feedback from the MSc and Executive Education programmes at Swansea University, and experiential learning from VBHC implementation in Wales, where national implementation efforts led to the formation of the Welsh Value in Health Centre and its national strategy in 2021.(4) Draft versions of the questionnaire were also reviewed by international VBHC leaders and practitioners to ensure relevance and clarity across different health system contexts.

Firstly, we wanted to test respondents’ confidence across four domains necessary for VBHC adoption: VBHC theory, outcomes measurement, costing methodology and the practical application of VBHC to healthcare transformation. These were then broken down further into questions about specific topics in each domain that are necessary for building VBHC infrastructure (e.g., costing and driving VBHC impact, the use of outcome data to support whole pathway redesign). We were also interested to ascertain gaps between interest in each area and confidence to implement, particularly between professional groups working within healthcare. The full questionnaire is available in Online Supplemental Appendix 1.

### Survey sample, data collection, and data analysis

Respondents were recruited initially using a snowball sampling technique. A request for participation was posted on social media platforms such as LinkedIn™, through professional and personal connections of the researchers, and through international dissemination channels, including the newsletter of the International Consortium for Health Outcomes Measurement (ICHOM). Leaders from several healthcare organisations globally distributed the survey information within their teams, while subsequent participants received the survey link from colleagues and professional contacts. The survey was open between January and June 2025. Results were analysed using IBM SPSS Statistics version 30.

## RESULTS

This approach resulted in 518 returned survey responses. Table 1 describes respondent characteristics. Over half reside in the UK and Ireland, with 19.7% from the Asia/Pacific region and 13.9% from mainland Europe. These were experienced health professionals, as 65% have been in their occupation for at least 10 years and 41% for more than 20 years. Respondents represented a wide range of professional roles. Approximately 32% of the respondents held clinical roles (17% physicians and 15% nurses or allied health), 12% held positions in the life sciences or in procurement, 11% were in executive roles or roles related to health finance, and 11% were operational managers in a health care setting.

**Table 1.** Respondent Characteristics.

| **Respondent Location** | **% (n)\*** |
| --- | --- |
| UK and Ireland | 51% (264) |
| Asia/Pacific Region | 20% (102) |
| Mainland Europe | 14% (72) |
| USA and Canada | 4% (22) |
| Middle East | 3% (14) |
| Central and South America | 2% (8) |
| Africa | <1% (2) |
| Did not answer | 7% (34) |
| **Respondent Occupation** | **% (n)\*** |
| Physician | 17% (88) |
| Nurse or Allied Health | 15% (80) |
| Life Sciences or Procurement | 12% (63) |
| Operational Manager | 11% (57) |
| Finance or Executive | 11% (55) |
| Other Occupational Groups* | 28% (143) |
| Did not answer | 6% (32) |

\*Only the largest occupational groups are reported separately. All others reported here.

### VBHC Challenges

Respondents were asked to identify their top VBHC implementation challenges from a pre-defined list. Of the 481 respondents who answered the question, difficulty implementing outcome measurement (62.4%) was the most frequently selected challenge, followed by conflicting priorities (57.7%), resistance to change (56.8%), lack of understanding of VBHC concepts (53.1%), insufficient resources (50.6%), and lack of access to trustworthy data (47.3%).

## Confidence and Interest in VBHC Domains

Respondents were next asked to rate their confidence in their knowledge of four broad areas of VBHC on a scale from 0-100, with 100 indicating the highest confidence level: VBHC Theory, Outcomes Measurement, Practical Application of VBHC and Costing. Respondents reported the highest confidence in VBHC Theory (median 70/100, interquartile range (IQR) 48-85), followed by Outcome Measurement (median 67/100, IQR 50-80), Practical Application of VBHC (median 55/100, IQR 31-75) and Costing (median 50/100, IQR 27-70). Responses spanned the full range from 0 to 100 across all four areas, indicating substantial variation in respondents’ confidence levels.

Gaps Between Level of Interest and Confidence in Ability to Enact Primary VBHC Implementation Domains Beyond the four broad VBHC areas, respondents rated both their interest in and confidence in their ability to implement 15 specific VBHC domains. Interest and confidence were measured using 0-100 point scales, with higher scores indicating greater interest or confidence. The results of these domains are displayed in Figure 1 (interest in VBHC domains) and Figure 2 (confidence in VBHC domains).

**Figure 1.**
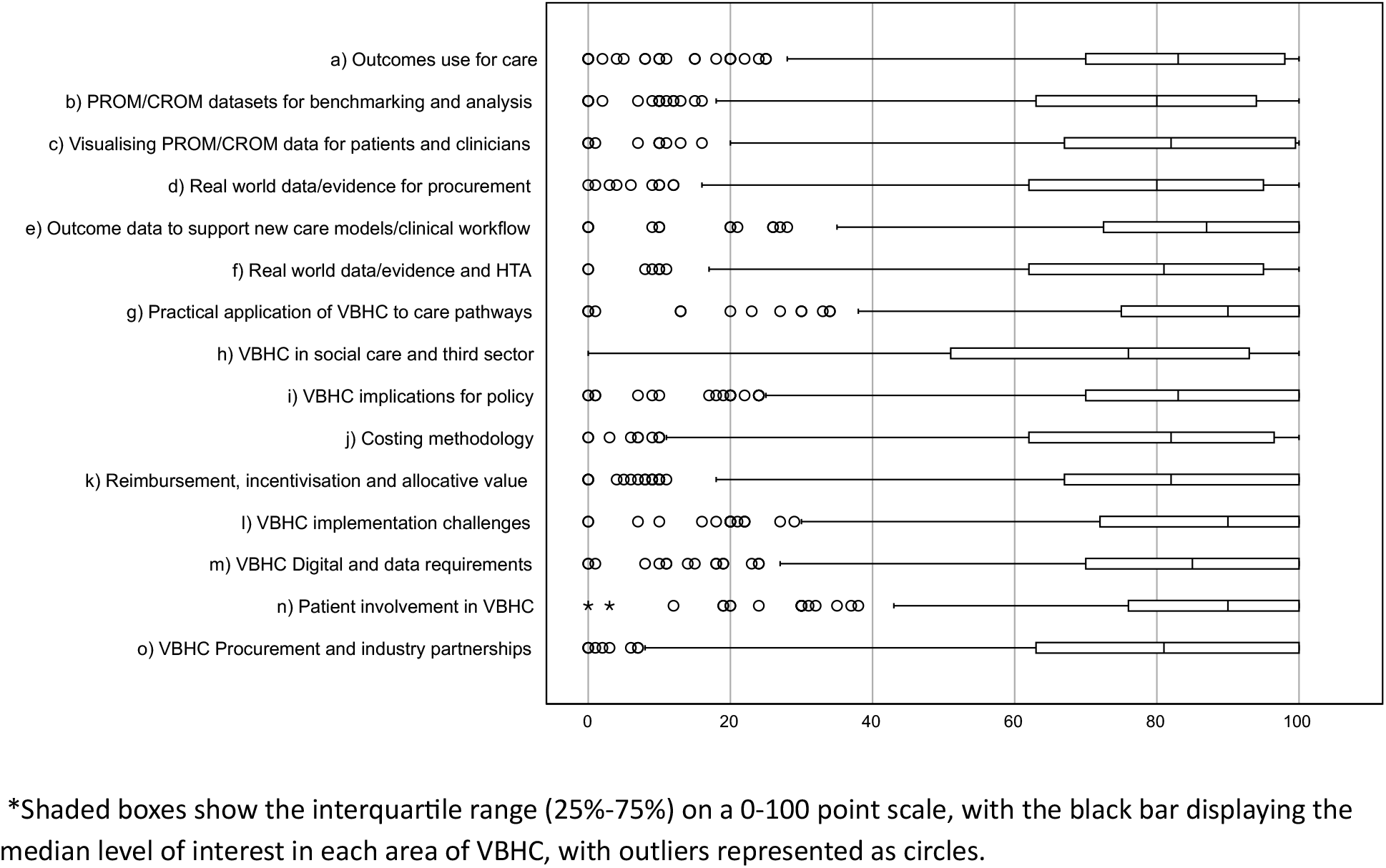
Boxplots indicating respondent level of interest in VBHC domains*.

**Figure 2.**
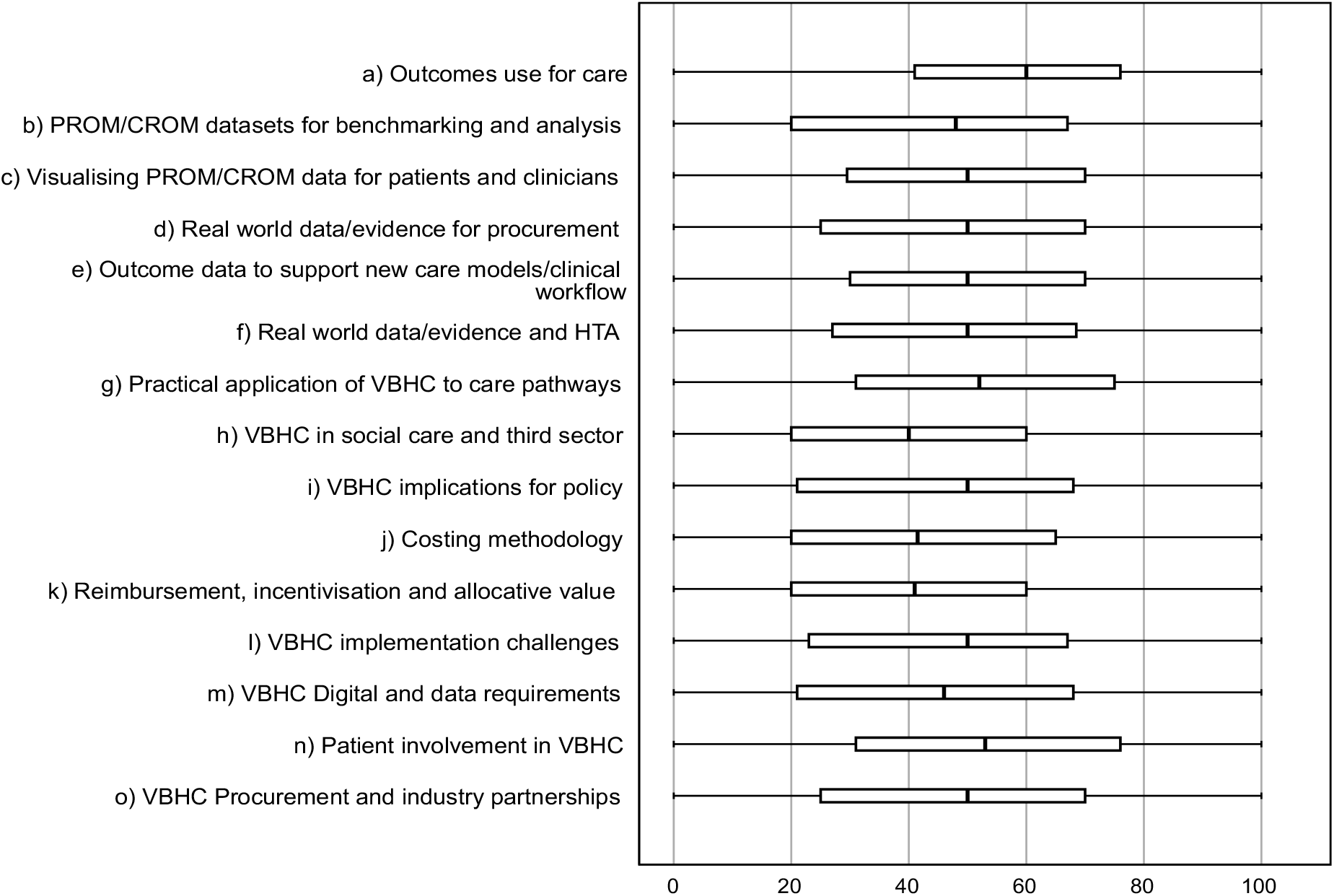
Boxplots indicating respondent level of confidence in VBHC domains*.

*Shaded boxes show the interquar3le range (25%-75%) on a 0-100 point scale, with the black bar displaying the median level of confidence in each area of VBHC.

Table 2 displays the resultant gaps between interest in a domain and confidence in being able to enact VBHC in that domain. Respondents reported strong interest in each VBHC domain. All median interest scores were above 80/100 except for “application of VBHC to social care and the third sector,” which fell just below (76/100). Domains with the highest interest scores were “overcoming implementation challenges for VBHC” and “patient involvement in VBHC” (median for each 90/100). While a few individuals indicated very low interest levels in the domains (0-20 points/100), these were outlier values, defined as those that fell outside 1.5x the interquartile range and marked with individual circles on the box and whisker plot.

**Table 2.** Gaps between interest and confidence in capability to enact VBHC domains*.

| VBHC Domain | Median Interest | Median Confidence | Median “gap” between interest and confidence |
| --- | --- | --- | --- |
| a) Outcomes use for care | 83 | 60 | 23 |
| b) PROM/CROM datasets for benchmarking and analysis | 80 | 48 | 32 |

| **VBHC Domain** | **Median Interest** | **Median Confidence** | **Median “gap” between interest and confidence** |
| --- | --- | --- | --- |
| c) Visualising PROM/CROM data for patients and clinicians | 82 | 50 | 32 |
| d) Real world data/evidence for procurement | 80 | 50 | 30 |
| e) Outcome data to support new care models/clinical workflow | 87 | 50 | 37 |
| f) Real world data/evidence and HTA | 81 | 50 | 31 |
| g) Practical application of VBHC to care pathways | 90 | 52 | 38 |
| h) VBHC in social care and 3<sup>rd</sup> sector | 76 | 40 | 36 |
| i) VBHC implications for policy | 83 | 50 | 33 |
| j) Costing methodology | 82 | 42 | 40 |
| k) Reimbursement, incentivisation, and allocative value | 82 | 41 | 41 |
| l) Overcoming implementation challenges | 90 | 50 | 40 |
| m) VBHC digital and data requirements | 85 | 46 | 39 |
| n) Patient involvement in VBHC | 90 | 53 | 37 |
| o) VBHC procurement and industry partnerships | 81 | 50 | 31 |

\*Measured on a 100-point scale. For each category of interest and confidence, the minimum value chosen by any respondent was 0 and the maximum value was 100.

In contrast, the highest median confidence score was for “confidence in the use of outcomes measurement for direct care, shared decision-making, and clinical decision-making” (60/100), followed by “patient involvement in VBHC (53/100) and “application of VBHC to care pathways” (52/100). All other confidence levels fell between 41-50/100. Each domain included respondents indicating “0” confidence and others marking “100” level of confidence, but unlike the self-reported interest levels, these low confidence scores were not just outliers, as indicated by the lower whisker bar (vs. circles) on the box and whisker plot.

The biggest gaps between interest and confidence were recorded for “reimbursement mechanisms, incentivisation, and allocative value (optimal use of resources across and between care pathways)” (median gap 41/100), “implementation challenges for VBHC” (median gap 40/100), and “costing methodology” (median gap 40/100). Most gaps had a median value in the range of 30-39/100, other than the domain for “confidence in the use of outcomes measurement for direct care, shared decision-making, and clinical decision-making,” with a median gap of just 23/100.

Given that professionals from different occupations may vary in level of interest and confidence, an ANOVA was run to compare ratings for each domain across the main occupational categories: 1. Doctors, 2. Allied Health Professionals/Nurses, 3. Managers, Executives and Finance, and 4. Procurement/Life Sciences. Only a few significant differences emerged. Procurement/Life Sciences professionals differed in a few areas. Their level of interest and confidence in “partnering with industry and value-based procurement” was unsurprisingly higher than other occupations, as was their confidence in “Real world data and evidence and health technology assessments.” Procurement and life sciences professionals also had statistically significant higher levels of interest and confidence compared to allied health professionals and nurses in the domains of “Digital and data requirements” and “Reimbursement mechanisms.”

### VBHC learning preferences and opportunities

The final question set on the survey was designed to understand what form of learning is preferable to these professionals and whether they already had accessed VBHC education and training (see Table 3). Respondents narrowly report a preference for in-person seminars (55%) and online masterclasses (54%) followed by online courses (50%) and case studies (49%). Fewer respondents preferred podcasts (29%) or white papers (24%).

**Table 3.**
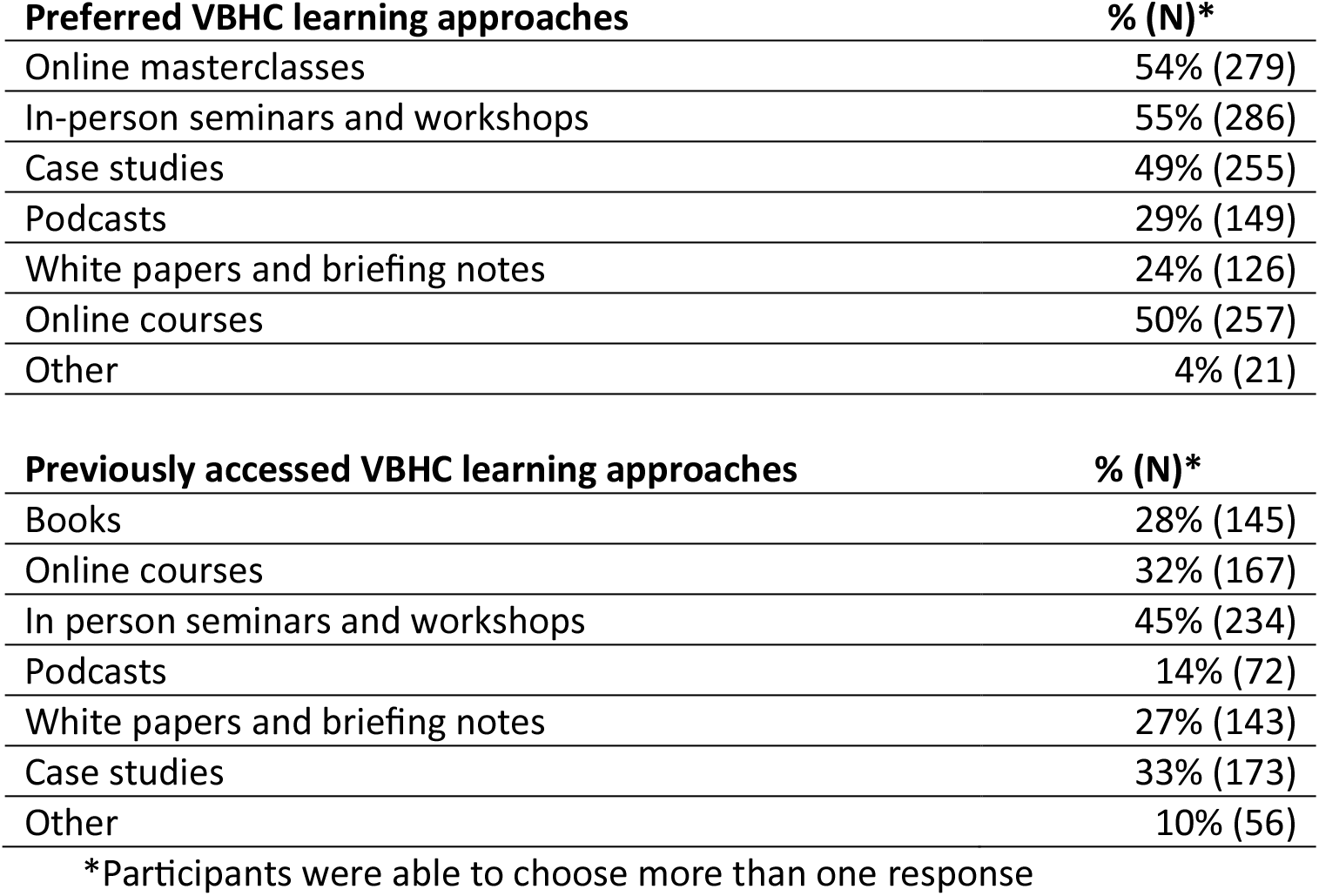
Preferred and Previously Accessed Approaches to VBHC Learning.

The most frequently accessed forms of learning were in-person seminars and workshops (45%), followed by case studies (33%) and online courses (32%). Self-directed methods such as white papers (27%), books (28%), and podcasts (14%) were reported less frequently.

## DISCUSSION

This exploratory, global needs assessment identified substantial gaps between health professionals’ interest inValue-Based Healthcare (VBHC) and their self-reported confidence to implement key VBHC domains in practice. Respondents expressed consistently high levels of interest across 15 VBHC domains, while confidence levels remained markedly lower, resulting in large interest–confidence gaps. These findings suggest that despite widespread engagement with VBHC concepts internationally, many health professionals do not yet feel fully equipped to translate these principles into routine practice.

Several findings merit attention. First, two of the largest competency gaps were observed in domains related to reimbursement mechanisms and costing methodologies. These areas are technically complex and appear addressed less prominently in VBHC curricula, as reflected in the lower confidence observed in these domains. In contrast, the comparatively smaller gap and higher confidence observed for outcomes measurement in direct care likely reflect its closer alignment with clinical practice and its central role in VBHC implementation and training. This is consistent with the emphasis placed on outcomes measurement in the VBHC literature.(2)(5) The predominantly clinical composition of the respondent population may also have contributed to this pattern, as procurement and life sciences professionals reported comparatively higher confidence in technical domains such as reimbursement mechanisms and value-based procurement. Together, these findings highlight the importance of strengthening technical VBHC capabilities within clinical teams while ensuring access to specialised organisational and analytical expertise to support implementation and system transformation.

Second, respondents reported substantial barriers to VBHC implementation, including difficulty implementing outcome measurement, competing priorities, and resistance to change. These challenges echo prior literature describing the organisational, cultural, and structural obstacles to VBHC adoption.(6) Notably, more than half of respondents also cited lack of understanding of VBHC concepts as a barrier, reinforcing the finding that high interest does not equate to implementation readiness. Together, these findings suggest that education should be integrated with organisational strategy, leadership support, and opportunities for advanced applied learning.

Third, respondents preferred interactive learning formats, such as in-person and online programmes and masterclasses, compared with self-directed modalities such as podcasts and white papers. The most frequently accessed learning formats broadly mirrored these preferences, suggesting alignment between preferred and available learning opportunities. This pattern indicates that health professionals may value opportunities to discuss concepts with peers and apply learning in practical contexts. However, the persistence of confidence gaps suggests that existing VBHC offerings may not adequately develop the specialised capabilities required for implementation, particularly in technical areas such as costing and reimbursement.

Fourth, differences across professional groups were limited but notable. Procurement and life sciences professionals reported higher levels of interest and confidence in domains such as value-based procurement, digital and data requirements, and reimbursement mechanisms. These findings highlight the importance of cross-disciplinary learning and closer alignment between technical, clinical, and organisational perspectives within VBHC education. Strengthening shared understanding across stakeholder groups can foster greater collaboration between clinical, industry, and health technology assessment perspectives in VBHC implementation.

The consistency of these findings across professional roles and multiple geographic regions suggest that these learning needs are widely shared. While health systems vary by context, common challenges in implementing VBHC were evident. This underscores a need for VBHC education and training programmes that emphasise advanced competencies, practical application and real-world examples.

## Limitations

This study has several limitations. The use of a non-probability, convenience sampling strategy limits the generalisability of findings as these respondents are likely to have greater interest in and experience with VBHC implementation compared to the broader population of health professionals. Self-reported confidence may not accurately reflect objective competence, and respondents may have different interpretation of these VBHC domains. In addition, while the survey captured a broad international sample, respondents were disproportionately based in the UK and Ireland. Despite these limitations, the large sample size, diversity of professional roles and consistency of findings support the value of this study as an exploratory needs assessment to inform workforce development and future research.

## CONCLUSION

This international exploratory survey demonstrates gaps between health professionals’ interest in value-based health care and their confidence to implement key VBHC elements in practice. While interest in VBHC concepts is high, particularly across foundational concepts such as outcomes measurement and patient involvement, confidence remains limited across most VBHC domains. These gaps align with commonly reported implementation challenges and reflect the VBHC learning most open available to and accessed by respondents.

Successful and sustainable VBHC initiatives are likely to require narrowing these gaps in confidence. While VBHC education must continue to address foundational principles, future training may benefit both from greater emphasis on technical capabilities, such as costing methodologies, data infrastructure, and value-based procurement, as well as on leadership and management capabilities, including strategies to overcome resistance to change.

By aligning VBHC education more closely with the practical implementation challenges faced by health professionals, health systems may better equip their workforce to translate value-based principles into meaningful improvements in care delivery and outcomes.

## Supporting information

Supplemental Appendix 1 Survey

## Data Availability

De-identified survey data are available from the corresponding author upon reasonable request.

## Acknowledgements

The authors thank the International Consortium for Health Outcomes Measurement (ICHOM) for supporting dissemination of the survey through its international network and newsletter. We also thank Jennifer Bright for reviewing a draft version of the manuscript and providing valuable feedback on the interpretation and presentation of the findings.

## Funding

No specific grant was received from any funding agency in the public, commercial or not-for-profit sectors for this research.

### Ethics approval

Ethical approval was granted by the Swansea University Research Ethics Committee. Participants were informed about the purpose of the study and completion of the survey was taken as implied consent to participate.

