## Supplemental Appendix 1 Survey for "A global cross-sectional survey of health professionals’ interest-confidence gaps in value-based health care implementation: a learning needs assessment"

qualtrics XM

---

Q1 Introduction: At the Swansea Academy for Value-based health and care, we want to know more about the interests and learning needs of professionals in health and care regarding Value-Based Healthcare. Your answers will help us develop masterclasses and educational programmes that are relevant and effective. Participation in this survey is anonymous. Completion and submission of this survey implies consent for your responses to be aggregated with others and used for research and publication. No personal data is collected.  What best describes your occupation and/or your perspective on VBHC?

- ○ Patient  (10)
- ○ Doctor  (1)
- ○ Allied Health Professional or Clinical Scientist  (2)
- ○ Nurse  (3)
- ○ C-suite/Executive of health provider  (5)
- ○ Operational manager in healthcare  (12)
- ○ Finance professional  (4)
- ○ Informatician  (8)
- ○ Procurement professional  (7)
- ○ Life science industry professional  (6)
- ○ Policy-maker  (9)
- ○ Academic  (11)
- ○ Other - please state below  (13)

________________________________________________________________

Q2 How many years have you been in this occupation?

- 0-5 (1)
- 5-10 (2)
- 10-20 (3)
- 20-30 (4)
- 30+ (5)

Q3 Where are you based?

- Nordic countries (1)
- UK and Ireland (2)
- Rest of Europe (3)
- Africa (4)
- Asia/Pacific (5)
- Canada (6)
- Central and South America (7)
- MIddle East (8)
- USA (9)

qualtrics XM

Q4 Please rate your confidence in the following value-based healthcare domains:

| | 0 10 20 30 40 50 60 70 80 90 100 |
|---|---|
| Outcomes measurement (1) | |
| Costing (2) | |
| VBHC theory (3) | |
| The practical application of VBHC to healthcare transformation (4) | |

Q5 What are the key challenges in implementing value-based healthcare in your setting? (Please select all that apply)

- ▢ Lack of understanding of value-based healthcare concepts (1)
- ▢ Difficulties with implementing outcome measurement (2)
- ▢ Conflicting priorities (3)
- ▢ Lack of trustworthy data (4)
- ▢ Insufficient resources (5)
- ▢ Resistance to change (6)
- ▢ Other (please specify below) (7)

________________________________________________

qualtrics XM

Q6 Outcome data can be used in multiple ways to support clinical, patient, administrative and policy decision-making in healthcare. Clinical outcome data (CROMs) are reported by clinicians and patient-reported outcome measures (PROMs) are reported by people receiving care. The latter may cover both symptom burden of a specific disease and overall quality of life. Please tell us how confident and interested you are personally in:  a) The use of outcome data for direct care/shared decision-making/clinical decision-making

| | 0 10 20 30 40 50 60 70 80 90 100 |
| --- | --- |
| Interest in the topic (2) | |
| Confidence in the topic (1) | |

b) Analysis of large PROM and CROM datasets for benchmarking and analysis, including predictive analytics and AI

| | 0 10 20 30 40 50 60 70 80 90 100 |
| --- | --- |
| Interest (2) | |
| Confidence in the topic (1) | |

c) Visualising PROM/CROM data for patients and clinicians

| | 0 10 20 30 40 50 60 70 80 90 100 |
| --- | --- |
| Interest (2) | |
| Confidence (1) | |

d) Real world data/evidence and its implications for procurement

| | 0 10 20 30 40 50 60 70 80 90 100 |
| --- | --- |
| Interest (2) | |
| Confidence (1) | |

qualtrics XM

e) Practical application of outcome data to support new models of care and clinical workflow (including as tools to support patient self-management)

| | 0 10 20 30 40 50 60 70 80 90 100 |
|---|---|
| Interest (2) | |
| Confidence (1) | |

f) Real world data/evidence and health technology assessments

| | 0 10 20 30 40 50 60 70 80 90 100 |
|---|---|
| Interest (2) | |
| Confidence (1) | |

Q7 Value-Based Healthcare impact requires the practical application of the principles to whole pathways of care. Please rate your confidence and your interest in this topic.

| | 0 10 20 30 40 50 60 70 80 90 100 |
|---|---|
| Interest (2) | |
| Confidence (1) | |

Q8 Please rate your confidence and your interest in the application of Value-Based Healthcare principles to social care and the third sector (NGOs and not-for-profit organisations)

| | 0 10 20 30 40 50 60 70 80 90 100 |
|---|---|
| Interest (2) | |
| Confidence (1) | |

Q9 Value-based Healthcare has key implications for policy-makers. Please rate your confidence and your interest in this topic.

| | 0 | 10 | 20 | 30 | 40 | 50 | 60 | 70 | 80 | 90 | 100 |
|---|---|---|---|---|---|---|---|---|---|---|---|
| Interest (2) | | | | | | | | | | | |
| Confidence (1) | | | | | | | | | | | |

Q10 Finance and human resources play a key role in Value-Based Healthcare, ensuring that we optimally use resources in healthcare systems. Please rate you interest and confidence in: a) costing methodology

| | 0 | 10 | 20 | 30 | 40 | 50 | 60 | 70 | 80 | 90 | 100 |
|---|---|---|---|---|---|---|---|---|---|---|---|
| Interest (2) | | | | | | | | | | | |
| Confidence (1) | | | | | | | | | | | |

b) Reimbursement mechanisms, incentivisation and allocative value (optimal use of resources across and between pathways of care)

| | 0 | 10 | 20 | 30 | 40 | 50 | 60 | 70 | 80 | 90 | 100 |
|---|---|---|---|---|---|---|---|---|---|---|---|
| Interest (2) | | | | | | | | | | | |
| Confidence (1) | | | | | | | | | | | |

Q11 There are significant challenges in the implementation of Value-Based Healthcare (including organisational culture, governance and change) but there are tried and tested ways of overcoming these. Please rate your confidence and interest in this topic.

| | 0 | 10 | 20 | 30 | 40 | 50 | 60 | 70 | 80 | 90 | 100 |
|---|---|---|---|---|---|---|---|---|---|---|---|
| Interest (2) | | | | | | | | | | | |
| Confidence (1) | | | | | | | | | | | |

Q12 Value-Based Healthcare is a data-driven discipline and digital technologies play a critical role in its implementation. Please rate your confidence and interest in the digital and data requirements for Value-Based Healthcare covering data standards, interoperability, visualisation, predictive analytics and AI.

| | 0 10 20 30 40 50 60 70 80 90 100 |
|---|---|
| Interest (2) | |
| Confidence (1) | |

Q13 Value-Based Healthcare is all about achieving the outcomes that matter to people. Patient involvement in Value-Based Healthcare at an individual and system level is therefore critical to success. Please rate your confidence and interest in this topic.

| | 0 10 20 30 40 50 60 70 80 90 100 |
|---|---|
| Interest (2) | |
| Confidence (1) | |

Q14 Value-Based Healthcare offers tremendous opportunities for the health sector and industry to work differently together towards more sustainable healthcare. Please rate your confidence and interest in Value-Based Procurement, Outcomes-Based agreements, partnering with industry

| | 0 10 20 30 40 50 60 70 80 90 100 |
|---|---|
| Interest (2) | |
| Confidence (1) | |

qualtrics XM

Q15 How do you like to learn? -Please select your preferences from these learning resources:

- ☐ Online Masterclasses (1)
- ☐ In person seminars/workshops (2)
- ☐ Case studies (3)
- ☐ Podcasts (4)
- ☐ White papers and briefing notes (5)
- ☐ Online courses (6)
- ☐ Other - please specify below (7)

________________________________________________

---

Q24 What learning resources for Value-Based Healthcare have you already accessed?

- ☐ Books (1)
- ☐ Online Courses (2)
- ☐ In person seminars/workshops (3)
- ☐ Podcasts (4)
- ☐ White papers and briefing notes (5)
- ☐ Case studies (6)
- ☐ Other - please specify below (7)

________________________________________________
